# Assessing the impact of the COVID-19 pandemic on routine childhood vaccination uptake in the Netherlands

**DOI:** 10.64898/2026.02.19.26346601

**Authors:** Joyce Pijpers, Manon Haverkate, Ruben van Gaalen, Susan Hahné, Hester de Melker, Susan van den Hof

## Abstract

**Background:** Initial reports from the Netherlands indicate a decline in routine childhood vaccination uptake during and after the COVID-19 pandemic, with emerging evidence of reduced parental vaccine confidence. This study aimed to evaluate the long-term impact of the COVID-19 pandemic on routine childhood vaccination uptake.

**Methods:** We conducted a retrospective nationwide cohort study including all children born in the Netherlands in 2016-2024. First-dose DTaP-IPV vaccination status by age six months was obtained from the national immunisation register. National trends in vaccination uptake across pre-pandemic, pandemic, and post-pandemic periods were assessed using interrupted time series analyses. To further assess the independent effect of the pandemic, a matched-sibling analysis compared vaccination uptake within families before, during and after the pandemic.

**Results:** Interrupted time series analyses showed significant immediate decreases in vaccination uptake both at the start and end of the pandemic, accompanied by a continuing downward trend during the pandemic (OR 0.984, 95%CI 0.982-0.985) that further declined after its end (OR 0.995, 95%CI 0.994-0.997). In the matched-sibling analysis children eligible during and after the pandemic had lower odds of being vaccinated (pandemic: OR 0.66, 95%CI 0.55-0.80; post-pandemic: OR 0.20, 95%CI 0.17-0.25) compared to their pre-pandemic siblings. Also, later birth order was associated with lower odds compared to first-born siblings (second-born: OR 0.42, 95%CI 0.37–0.48).

**Conclusions:** Both analyses indicate a negative impact of the COVID-19 pandemic on parental vaccination decisions, which may reflect lingering pandemic effects or new post-pandemic factors, highlighting the need for further research into the drivers of vaccination uptake changes in the post-pandemic era.

## Introduction

The COVID-19 pandemic has had major impact on all aspects of healthcare worldwide [1]. In the Netherlands, the emergence of COVID-19 in February 2020 led to the rapid implementation of widespread public health measures. The Dutch government introduced a lockdown mid-March 2020, which included closures of schools, restaurants, and non-essential shops. Alternating periods of stricter and more relaxed restrictions followed, before most measures were lifted early 2022 [2].

Although infant vaccination schedules were largely maintained, appointments for older children and adolescents were occasionally postponed or rescheduled. Despite catch-up campaigns, early data from the Netherlands indicated that the vaccination coverage for infants scheduled to receive their first measles, mumps, rubella (MMR1) vaccination between March and September 2020, was estimated to be 1-2 percentage points lower compared to the coverage among children born one year earlier [3]. Importantly, a decreasing trend in first-dose MMR and ≥3-dose DTaP-IPV vaccination coverage (by age 2 years) was already apparent before the pandemic. Coverage, which had been consistently above 95% for the 2005–2011 birth cohorts, dropped to 92.6% for DTaP-IPV and 92.9% for MMR in cohort 2015. After a slight recovery to 93.1% (DTaP-IPV) and 93.3% (MMR) for the 2018 birth cohort, coverage declined again, reaching 87.9% and 87.7% for MMR and DTaP-IPV, respectively, in the 2022 birth cohort [4] (see supplementary figure S1). Notably, from 2022 onwards, a new policy requiring explicit parental informed consent for vaccination registration was implemented in the Netherlands. Consequently, children vaccinated without parental consent for registration are recorded as unvaccinated, leading to a underestimation of coverage in recent years.

In addition to logistical barriers, the pandemic coincided with an unprecedented spread of mis- and disinformation and scepticism surrounding the SARS-CoV-2 virus, the public health measures and vaccines, particularly during the COVID-19 vaccine rollout [5]. Emerging evidence suggests that this infodemic has undermined trust not only in COVID-19 vaccines, but also in established childhood immunisation programmes. For example, UNICEF reported a worldwide decline in confidence in the importance of childhood vaccination, including the Netherlands (−10 percentage points in 2022 compared to 2019) [6]. Similarly, surveys in 2022 and 2023 indicated that Dutch parents have become slightly more negative towards routine childhood vaccines compared to a decade earlier [7, 8]. These findings suggest the pandemic’s impact may extend beyond temporary service disruptions towards a sustained decrease in vaccine uptake.

Therefore, this study aimed to assess this impact by using two complementary approaches. First, we performed an interrupted time series analysis (ITS) to assess changes in population level vaccination trends and to determine whether the rate of decline in vaccine uptake changed at the start and at the end of the pandemic. Second, we use a matched-sibling analysis to compare siblings within the same family, thereby assessing the effect of the pandemic while controlling for family-specific factors that may influence vaccination decisions. Together, these analyses provide a comprehensive assessment of the pandemic’s impact on routine childhood vaccination in the Netherlands.

## Methods

### Study setting and population

This retrospective registry study was conducted using linked national data within the secure remote access environment of Statistics Netherlands (Centraal Bureau voor de Statistiek, CBS). We included children born 2016–2024, residing in the Netherlands on 31-12-2024, as registered in the personal records database. Using national registries on migration, we excluded children who were not living in the Netherlands during the pre-defined age window for eligibility for first-dose DTaP-IPV vaccination (2-6 months of age). Children were assigned to one of three pandemic periods based on their date of birth and the corresponding age of eligibility for their first-dose DTaP-IPV vaccination (Table 1). Pandemic periods were defined based on COVID-19-related public health measures in the Netherlands. The relaxation of pandemic measures coincided in time with the introduction of informed consent of registration of vaccination in the national vaccination register Praeventis [9]. National data indicate that on average 1.7% of parents did not provide consent for primary series DTaP-IPV vaccination registration.

**Table 1.** Definitions of pandemic periods and corresponding birth cohorts.

| Period | Definition | Birth cohort |
| --- | --- | --- |
| Pre-pandemic | Eligible for first-dose DTaP-IPV before implementation of pandemic response measures in March 2020 | 1 January 2016 – 31 December 2019 |
| Pandemic | Eligible for vaccination during the period of COVID-19 measures and prior to the introduction of informed consent for national vaccination registration (March 2020 – January 2022) | 1 January 2020 – 30 September 2021 |
| Post-pandemic | Eligible for vaccination after the relaxation of pandemic measures and following the introduction of the informed consent policy in January 2022 | 1 October 2021 – 31 December 2024 |

### Outcome measure

The primary outcome was having received the first-dose DTaP-IPV vaccination at the age of 6 months. The first dose is scheduled at 2 months of age for children born before 2020. In 2020 maternal pertussis vaccination was introduced, and children of vaccinated mothers are scheduled to receive the first dose at 3 months. Otherwise, the first dose is given at 2 months of age. Individual level vaccination status at 6 months of age was retrieved from Praeventis on 28 August 2025.

### Interrupted time series analysis

We used an interrupted time series (ITS) design to assess changes in first-dose DTaP-IPV vaccination uptake. Two interruptions were defined, corresponding to the transitions between the previously defined ‘pandemic periods.’ ITS is a quasi-experimental method for evaluating changes in outcomes following distinct intervention points or events [10]. We used individual-level data and segmented regression models to estimate changes in the probability of receiving the first scheduled DTaP-IPV vaccination associated with each interruption, capturing both immediate (level) and gradual (slope) changes over time. A binomial logistic regression model was used. Results are presented as odds ratios with 95% confidence intervals, where each period is compared to the immediately preceding period (i.e., the pandemic period is compared to the pre-pandemic period, and the post-pandemic period is compared to the pandemic period). Average monthly model predictions were used to calculate the absolute level changes (i.e., percentage point changes in vaccination uptake).

First, a simple model was created, including birth year and month, interruption 1 and 2 (level changes) and time since interruption 1 and 2 (slope changes). In the second model we extended the first model by adjusting for sociodemographic variables, including birth order, migration status, maternal education level, household income, and urbanisation level based on the residential neighbourhood. For descriptive statistics, we also included the variable country of origin. Because some categories of ‘country of origin’ and ‘migration status’ (e.g. ‘Dutch origin’ and ‘country of origin: The Netherlands’) overlapped completely, including both variables in the model would result in multicollinearity. The reference year for all sociodemographic variables was 2024. A more detailed description of the sociodemographic variables can be found in supplementary table S2.

### Matched-sibling analysis

We performed a matched-sibling analysis to improve the possibilities for causal inference by accounting for unmeasured family-level confounders. We included families with at least two children, with all children born in 2016-2024. Families in which any child was living outside the Netherlands at the time of first-dose DTaP-IPV vaccination eligibility were excluded. In the case of multiples, we included only the first child in the register, as vaccination decisions for multiples are typically made collectively and reflect a single family-level decision.

We employed conditional logistic regression to compare vaccination uptake among siblings within the same family, effectively matching on shared family-level factors. Birth order was included as an independent variable, as previous studies have shown that later-born children are at higher risk of incomplete or delayed vaccination [11]. We aimed to investigate whether this association persisted or changed during and after the pandemic. Therefore, we first constructed a model that included birth order and pandemic period as independent variables. In a second model, we added an interaction term between pandemic period and birth order to assess whether the effect of birth order on vaccination uptake differed before, during, and after the pandemic. In accordance with the principles of conditional logistic regression, only discordant families - those in which siblings differed in outcome - contributed to the estimation of model parameters. Furthermore, for the estimation of the effect of ‘pandemic period,’ only discordant families in which siblings were eligible for vaccination in different pandemic periods (i.e., before, during, or after the pandemic) contributed information to these parameter estimates [12]. Descriptive family-level characteristics were reported to provide context for generalizability of the findings.

### Sensitivity analyses

As a sensitivity analysis, we repeated the conditional logistic regression analysis restricted to families with two children to assess consistency and generalizability across different family sizes. This approach allows for a straightforward comparison between first- and second-born siblings and reduces potential heterogeneity associated with larger family sizes, which may differ in parental characteristics or vaccination behaviours [13]. In this subgroup, by definition, first-born children were always eligible pre-pandemic and second-born children during or after the pandemic. As a result, birth order and pandemic period were perfectly overlapping, and only the main effects model (model 1) was estimated, as independent estimation of interaction effects was not possible.

To assess the potential impact of misclassification due to incomplete parental consent for national vaccination registration after January 2022, we conducted a sensitivity analysis in which we reclassified a proportion of unvaccinated children eligible in the post-pandemic/informed consent period (2021–2024) as vaccinated. Specifically, since the 1.7% non-consent rate refers to the vaccinated population, we calculated the number of unvaccinated children to be reclassified as vaccinated using the formula: (number of vaccinated children in the subset / 98.3) × 1.7. For the ITS analysis, we did not apply this correction, as adjusting for non-consent would only shift the absolute coverage level in the post-pandemic period upward, without affecting the underlying slope over time. Therefore, the potential impact of incomplete consent on the ITS results is addressed in the interpretation of our findings rather than through direct adjustment of the estimates.

All analyses were performed in R version 4.4.3[14].

## Results

The total study population included 1,375,514 children of whom 1,284,440 (93%) had received a first-dose DTaP-IPV vaccination at 6 months of age (Table 2). Vaccination coverage was highest in the pre-pandemic period and declined slightly during and after the pandemic. Uptake was generally higher among first- and second-born children compared to later born children, those with Dutch origin compared to non-Dutch origin, higher maternal education level and higher household income versus lower maternal education level and lower household income. Coverage also varied by country of origin and level of urbanisation.

**Table 2.** First-dose DTaP-IPV vaccination uptake at age 6 months by descriptive sociodemographic characteristics, The Netherlands, children born 2016-2024 (n = 1,375,514)

| Characteristics | Total population |  | Vaccinated population |  |
| --- | --- | --- | --- | --- |
|  | N | Column % | N | Row % |
| Total number of children | 1,375,514 | 100 | 1,284,440 | 93 |
| Pandemic period |  |  |  |  |
| Pre-pandemic | 589,003 | 43 | 562,205 | 95 |
| Pandemic | 277,223 | 20 | 260,742 | 94 |
| Post-pandemic | 509,288 | 37 | 461,493 | 91 |
| Birth order |  |  |  |  |
| First born | 660,026 | 48 | 624,849 | 95 |
| Second born | 523,562 | 38 | 491,982 | 94 |
| Third born | 156,408 | 11 | 140,233 | 90 |
| Fourth or later born | 35,518 | 3 | 27,376 | 77 |
| Year of birth |  |  |  |  |
| 2016 | 144,177 | 10 | 137,232 | 95 |
| 2017 | 145,791 | 11 | 139,035 | 95 |
| 2018 | 147,766 | 11 | 141,065 | 95 |
| 2019 | 151,269 | 11 | 144,873 | 96 |
| 2020 | 153,090 | 11 | 144,413 | 94 |
| 2021 | 165,515 | 12 | 154,694 | 93 |
| 2022 | 155,927 | 11 | 142,905 | 92 |
| 2023 | 154,422 | 11 | 139,444 | 90 |
| 2024 | 157,557 | 11 | 140,779 | 89 |
| Family size |  |  |  |  |
| One child | 324,162 | 24 | 303,861 | 94 |
| Two children | 700,061 | 51 | 666,334 | 95 |
| Three children | 275,127 | 20 | 252,911 | 92 |
| Four or more children | 76,164 | 6 | 61,334 | 81 |
| Migration status |  |  |  |  |
| Dutch origin | 830,040 | 60 | 786,450 | 95 |
| Migrant or child of first generation migrant(s) | 336,263 | 24 | 312,183 | 93 |
| Child of second generation migrant(s) | 209,211 | 15 | 185,807 | 89 |
| Country of origin <sup>a</sup> |  |  |  |  |
| The Netherlands | 830,040 | 60 | 786,450 | 95 |
| Europe (excl. The Netherlands) | 152,629 | 11 | 143,539 | 94 |
| Morocco | 59,793 | 4 | 44,198 | 74 |
| Turkey | 51,613 | 4 | 44,796 | 87 |
| Suriname | 41,732 | 3 | 38,607 | 93 |
| The Dutch Caribbean | 26,725 | 2 | 24,514 | 92 |
| Indonesia | 35,855 | 3 | 34,120 | 95 |
| Other, America/Oceania | 39,738 | 3 | 37,548 | 94 |
| Other, Africa | 47,652 | 3 | 44,793 | 94 |
| Other, Asia | 89,737 | 7 | 85,875 | 96 |
| Maternal education level |  |  |  |  |
| High | 658,273 | 48 | 624,970 | 95 |
| Not high | 717,241 | 52 | 659,470 | 92 |
| Household income <sup>b</sup> |  |  |  |  |
| First quartile | 148,969 | 11 | 131,572 | 88 |
| Second quartile | 343,537 | 25 | 310,967 | 91 |
| Third quartile | 502,183 | 37 | 476,197 | 95 |
| Fourth quartile | 375,579 | 27 | 361,292 | 96 |
| Unknown | 5,246 | <1 | 4,412 | 84 |
| Level of urbanisation |  |  |  |  |
| Not urbanised | 216,838 | 16 | 200,647 | 93 |
| Hardly urbanised | 242,206 | 18 | 228,561 | 94 |
| Moderately urbanised | 264,172 | 19 | 249,176 | 94 |
| Strongly urbanised | 351,509 | 26 | 329,207 | 94 |
| Extremely urbanised | 300,313 | 22 | 276,409 | 92 |
| Unknown | 476 | <1 | 440 | 92 |

<sup>a</sup> The country of origin is determined based on the country of birth of the child included in the study, and the countries/countries of birth of its parents and grandparents. <sup>b</sup> Standardised disposable income of the household. If parents of children were living in separate households, the average SDI of the two households was taken. Quartiles (percentiles) were calculated by CBS based on the complete Dutch population. All reported tables exclude frequencies below ten to avoid personally identifiable information.

### Interrupted time series analysis

The basic ITS model just taking into account year and month of birth showed an initial significant drop in vaccination uptake after the start of the pandemic with an OR of 0.777 (95% CI 0.746–0.809). This corresponds to an absolute level change of −1.12 percent points. Simultaneously, the slightly increasing trend in vaccination uptake that was visible from 2016-2019 (OR 1.004, 95% CI 1.003–1.004), significantly decreased after the start of the pandemic (OR 0.985, 95% CI 0.983–0.988). At the end of the pandemic, a significant level change was observed (OR 0.868, 95% CI 0.836–0.900), corresponding to an absolute level change of −1.00 percent points. No significant post-pandemic slope change was found compared to the pandemic period (OR 0.999, 95% CI 0.996–1.002).

After adjusting for sociodemographic factors, similar statistically significant decreases in vaccination uptake were seen directly after both interruptions (OR 0.765 [95% CI 0.735–0.798] and −0.896 [95% CI 0.863–0.930] at the start and end of the pandemic, as can be seen in Figure 1. These correspond to level changes of −1.15 and −0.66 percent points at the start and end of the pandemic, respectively. After the start of the pandemic, a significant slope change was found (OR 0.984, 95% CI 0.981–0.986) compared to pre-pandemic, indicating a decreasing trend in vaccination uptake. Opposed to model 1, in the adjusted model we did find a statistically significant slope change after the end of the pandemic compared to during the pandemic, with an OR of 0.995 (95% CI 0.993–0.998), indicating that the decreasing trend in vaccination uptake during the pandemic declined even faster after the end of the pandemic. Furthermore, when comparing the vaccination uptake as presented in Table 2 to the adjusted estimates of the covariates in model 2, similar trends in vaccination uptake are seen across specific subgroups. More details can be found in Table 3.

**Table 3.** Interrupted time series model results for first-dose DTaP-IPV vaccination uptake at age 6 months: unadjusted (model 1, n = 1,375,514) and adjusted for sociodemographic factors (model 2, n = 1,369,801)

|  | Model 1 (unadjusted) |  |  | Model 2 (adjusted) |  |  |
| --- | --- | --- | --- | --- | --- | --- |
|  | OR | 95% CI | p value | OR | 95% CI | p value |
| Slope (birth month) | 1.004 | 1.003 - 1.004 | <0.001 | 1.006 | 1.005 - 1.007 | <0.001 |
| Level change start pandemic <sup>a</sup> | 0.777 | 0.746 - 0.809 | <0.001 | 0.765 | 0.735 - 0.798 | <0.001 |
| Slope change start pandemic <sup>a</sup> | 0.985 | 0.983 - 0.988 | <0.001 | 0.984 | 0.981 - 0.986 | <0.001 |
| Level change end pandemic/start informed consent <sup>b</sup> | 0.868 | 0.836 - 0.900 | <0.001 | 0.896 | 0.863 - 0.930 | <0.001 |
| Slope change end pandemic/start informed consent <sup>b</sup> | 0.999 | 0.996 - 1.002 | 0.558 | 0.995 | 0.993 - 0.998 | 0.001 |
| Birth order |  |  |  |  |  |  |
| First born |  |  |  | ref | ref |  |
| Second born |  |  |  | 0.907 | 0.892 - 0.921 | <0.001 |
| Third born |  |  |  | 0.552 | 0.541 - 0.563 | <0.001 |
| Fourth born or later |  |  |  | 0.256 | 0.249 - 0.264 | <0.001 |
| Migration status |  |  |  |  |  |  |
| Dutch origin |  |  |  | ref | ref |  |
| Child of first generation migrant(s) |  |  |  | 0.926 | 0.909 - 0.943 | <0.001 |
| Child of second generation migrant(s) |  |  |  | 0.467 | 0.459 - 0.475 | <0.001 |
| Migrant |  |  |  | 0.306 | 0.278 - 0.336 | <0.001 |
| Maternal education level |  |  |  |  |  |  |
| High |  |  |  | ref | ref | <0.001 |
| Not high |  |  |  | 0.864 | 0.850 - 0.877 | <0.001 |
| Household income <sup>c</sup> |  |  |  |  |  |  |
| Fourth quartile |  |  |  | ref | ref |  |
| Third quartile |  |  |  | 0.700 | 0.685 - 0.715 | <0.001 |
| Second quartile |  |  |  | 0.395 | 0.387 - 0.404 | <0.001 |
| First quartile |  |  |  | 0.347 | 0.339 - 0.356 | <0.001 |
| Level of urbanisation |  |  |  |  |  |  |
| Not urbanised |  |  |  | ref | ref |  |
| Hardly urbanised |  |  |  | 1.352 | 1.320 - 1.385 | <0.001 |
| Moderately urbanised |  |  |  | 1.418 | 1.385 - 1.452 | <0.001 |
| Strongly urbanised |  |  |  | 1.357 | 1.327 - 1.388 | <0.001 |
| Extremely urbanised |  |  |  | 1.151 | 1.125 - 1.178 | <0.001 |

<sup>a</sup> Reference period: pre-pandemic. <sup>b</sup> Reference period: pandemic. <sup>c</sup> Standardised disposable income of the household. If parents of children were living in separate households, the average SDI of the two households was taken. Quartiles (percentiles) were calculated by CBS based on the complete Dutch population. All reported tables exclude frequencies below ten to avoid personally identifiable information.

**Figure 1.**
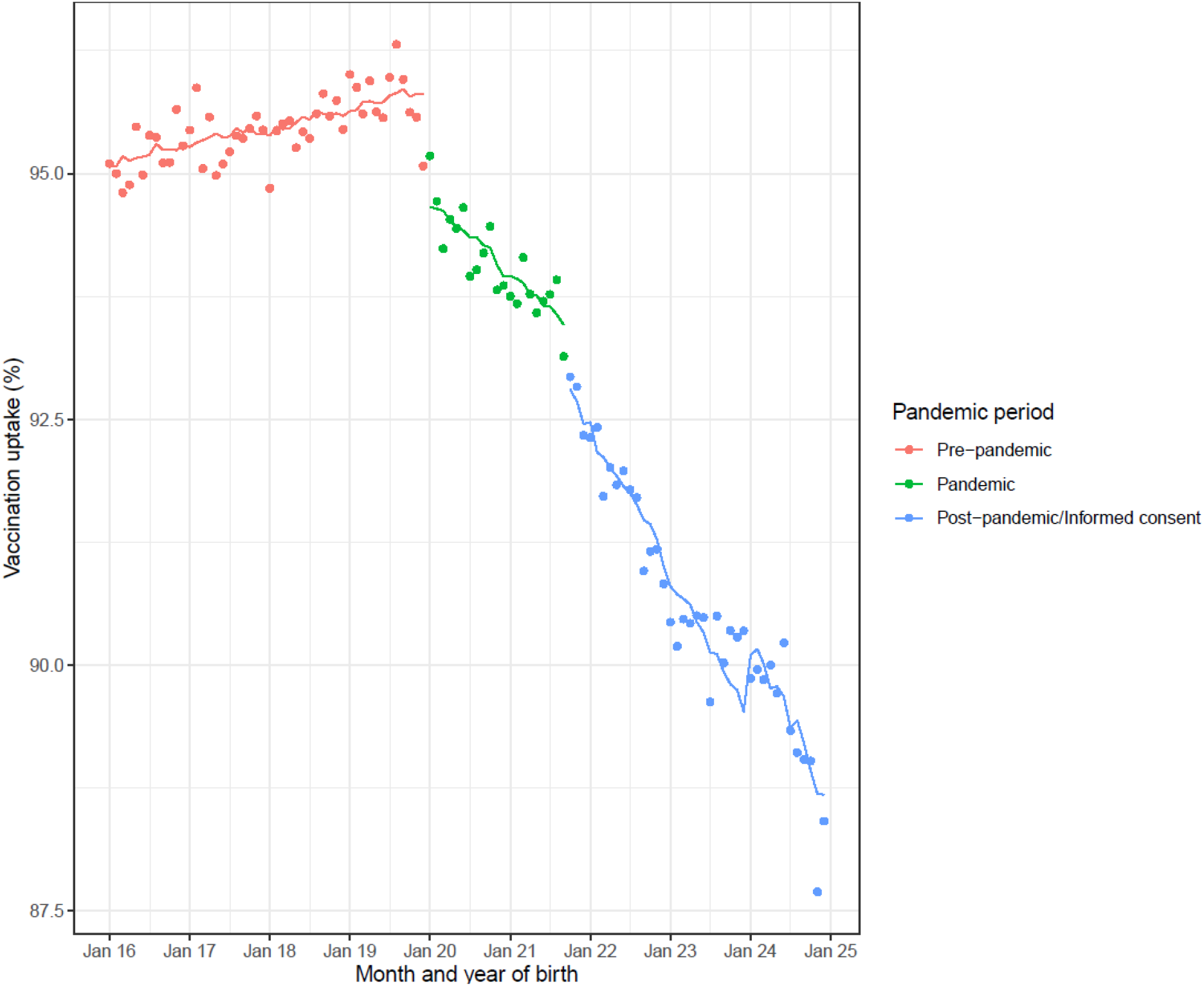
Vaccination coverage DTaP-IPV at age 6 months by month of birth and pandemic period, with fitted regression lines (model 2 - adjusted model). The figure displays month and year of birth: children born in January 2020 were eligible for first dose DTaP-IPV vaccination in March 2020 (start of the pandemic), and children born in October 2021 were eligible in January 2022 (end of the pandemic/start of informed consent). Points: observed data. Lines: model prediction. N.B. the model predictions (lines) are not linear, as they are based on the average predicted vaccination uptake per month, e.g., predictions are made on an individual level, and then an average predicted vaccination uptake per month is calculated and shown in the Figure.

### Matched-sibling analysis

Table 4 presents the descriptive characteristics of concordant and discordant families included in the matched-sibling analysis. The study population included 708,059 children within 326,981 families. The majority of families was concordant for vaccination status, with 301,465 families (92%) having all eligible children vaccinated and 13,164 families (4%) having all unvaccinated. The remaining 12,352 families (4%) were discordant, meaning siblings differed in vaccination status and thus contributed information to the conditional logistic regression. Most families had two children (86% of concordant-all-vaccinated, 74% of concordant-all-unvaccinated, and 68% of discordant families). Discordant families were more often of non-Dutch origin, had a lower maternal education level, and lived in more urbanised areas compared to concordant families, particularly compared to those in which all children were vaccinated.

**Table 4.** Family-level characteristics by sibling vaccination concordance for first-dose DTaP-IPV vaccination uptake at age 6 months, families with at least two children born 2016-2024 (n = 326,981)

|  | Concordant families |  |  |  | Discordant families |  |
| --- | --- | --- | --- | --- | --- | --- |
|  | All vaccinated |  | All unvaccinated |  |  |  |
|  | N | Column % | N | Column % | N | Column % |
| Total | 301,465 |  | 13,164 |  | 12,352 |  |
| Family size |  |  |  |  |  |  |
| Two children | 259,145 | 86 | 9,695 | 74 | 8,408 | 68 |
| Three children | 39,573 | 13 | 2,780 | 21 | 3,341 | 27 |
| Four or more children | 2,747 | 1 | 689 | 5 | 603 | 5 |
| Migration status |  |  |  |  |  |  |
| Dutch origin | 199,511 | 66 | 6,649 | 51 | 4,755 | 38 |
| Migrant or child of first generation migrant(s) | 58,756 | 19 | 2,939 | 22 | 3,782 | 31 |
| Child of second generation migrant(s) | 43,198 | 14 | 3,576 | 27 | 3,815 | 31 |
| Country of origin <sup>a</sup> |  |  |  |  |  |  |
| The Netherlands | 199,511 | 66 | 6,649 | 51 | 4,755 | 38 |
| Europe (excl. The Netherlands) | 30,346 | 10 | 1,126 | 9 | 1,184 | 10 |
| Morocco | 8,142 | 3 | 2,454 | 19 | 2,419 | 20 |
| Turkey | 972 | 7 | 972 | 7 | 1,437 | 12 |
| Suriname | 7,752 | 3 | 343 | 3 | 516 | 4 |
| The Dutch Caribbean | 4,837 | 2 | 263 | 2 | 344 | 3 |
| Indonesia | 7,869 | 3 | 223 | 2 | 240 | 2 |
| Other, America/Oceania | 8,179 | 3 | 287 | 2 | 273 | 2 |
| Other, Africa | 8,825 | 3 | 347 | 3 | 527 | 4 |
| Other, Asia | 16,687 | 6 | 500 | 4 | 657 | 5 |
| Maternal education level |  |  |  |  |  |  |
| High | 159,949 | 53 | 4,820 | 37 | 4,487 | 36 |
| Not high | 141,516 | 47 | 8,344 | 63 | 7,865 | 64 |
| Household income <sup>b</sup> |  |  |  |  |  |  |
| First quartile | 24,580 | 8 | 2,441 | 19 | 2,260 | 18 |
| Second quartile | 70,417 | 23 | 4,930 | 37 | 4,269 | 35 |
| Third quartile | 121,283 | 40 | 3,877 | 29 | 3,836 | 31 |
| Fourth quartile | 84,624 | 28 | 1,831 | 14 | 1,913 | 15 |
| Unknown | 561 | <1 | 85 | <1 | 74 | <1 |
| Level of urbanisation |  |  |  |  |  |  |
| Not urbanised | 50,879 | 17 | 2,599 | 20 | 1,564 | 13 |
| Hardly urbanised | 57,642 | 19 | 2,080 | 16 | 1,685 | 14 |
| Moderately urbanised | 60,669 | 20 | 2,239 | 17 | 2,010 | 16 |
| Strongly urbanised | 75,398 | 25 | 3,121 | 24 | 3,340 | 27 |
| Extremely urbanised | 56,776 | 19 | 3,121 | 24 | 3,747 | 30 |
| Unknown | 101 | <1 | <10 | <1 | <10 | <1 |

<sup>a</sup> The country of origin is determined based on the country of birth of the child included in the study, and the countries/countries of birth of its parents and grandparents. This characteristics table provides context for understanding the analytic sample; however, by design, the within-family analysis controls for stable family-level factors, and thus these differences do not confound the estimated effects within discordant families. All reported tables exclude frequencies below ten to avoid personally identifiable information.

Table 5 presents the results of the matched-sibling conditional logistic regression models. In Model 1, both pandemic period and birth order were independently associated with vaccination uptake within discordant families. Children who became eligible for vaccination during the pandemic had lower odds of receiving the first-dose DTaP-IPV vaccination compared to their siblings eligible before the pandemic (OR 0.50, 95% CI 0.44–0.55), and this effect was even more pronounced for those eligible after the pandemic (OR 0.15, 95% CI 0.13–0.18). Birth order also showed a strong association with vaccination uptake: compared to first-born siblings, the odds of vaccination were lower for second-born (OR 0.32, 95% CI 0.29–0.35), third-born (OR 0.14, 95% CI 0.12–0.16), and fourth or later-born children (OR 0.08, 95% CI 0.06–0.10).

**Table 5.** Matched-sibling conditional logistic regression model for first-dose DTaP-IPV vaccination uptake at age 6 months, children born 2016-2024 within families with at least two children (n = 708,059)

|  | Model 1 |  |  | Model 2 |  |  |
| --- | --- | --- | --- | --- | --- | --- |
|  | OR | 95% CI | p value | OR | 95% CI | p value |
| Pre-pandemic (ref) | ref | ref | Ref | ref | ref | ref |
| Pandemic | 0.50 | 0.44-0.55 | <0.001 | 0.66 | 0.55-0.80 | <0.001 |
| Post pandemic | 0.15 | 0.13-0.18 | <0.001 | 0.20 | 0.17-0.25 | <0.001 |
| First born (ref) | ref | ref | Ref | ref | ref | ref |
| Second born | 0.32 | 0.29-0.35 | <0.001 | 0.42 | 0.37-0.48 | <0.001 |
| Third born | 0.14 | 0.12-0.16 | <0.001 | 0.19 | 0.10-0.35 | <0.001 |
| Fourth or later born | 0.08 | 0.06-0.10 | <0.001 | 0.07 | 0.00-3.75 | 0.193 |
| pandemic* second born |  |  |  | 0.53 | 0.42-0.68 | <0.001 |
| post pandemic * second born |  |  |  | 0.66 | 0.55-0.79 | <0.001 |
| pandemic * third born |  |  |  | 0.64 | 0.33-1.24 | 0.182 |
| post pandemic * third born |  |  |  | 0.60 | 0.32-1.13 | 0.117 |
| pandemic * fourth or later born |  |  |  | 0.43 | 0.01-24.65 | 0.682 |
| post pandemic * fourth or later born |  |  |  | 0.94 | 0.02-47.86 | 0.973 |

In Model 2, the effect of birth order on vaccination uptake was modified by the pandemic period. For second-born children, the interaction terms were statistically significant, indicating lower odds of vaccination during and after the pandemic compared to first-borns (OR 0.53, 95% CI 0.42–0.68 and OR 0.66, 95% CI 0.55–0.79 for pandemic and post-pandemic, respectively). For third-born and fourth or later-born children, the interaction terms were not statistically significant. The total effects of birth order and pandemic period on vaccination uptake, calculated relative to first-born children eligible for vaccination pre-pandemic, are presented in Supplementary Table S3.

### Sensitivity analysis

We first repeated the matched-sibling conditional logistic regression analysis (model 1), restricting the sample to families with exactly two children (n = 554,496; see Supplementary Table S4). Results were nearly identical to those of the main analysis.

Additionally, we conducted a sensitivity analysis to address incomplete parental consent in the post-pandemic/informed consent period by reclassifying 16% of unvaccinated children as vaccinated. This proportion was calculated to reflect the estimated level of non-consent. The results remained broadly consistent with the primary analysis, although the effect sizes for birth order were smaller and the interaction between being second-born and the post-pandemic period was not statistically significant (see Supplementary Tables S5 and S6).

## Discussion

This study assessed the impact of the COVID-19 pandemic on routine childhood vaccination uptake using two complementary approaches. The ITS showed that the slight increase in first-dose DTaP-IPV uptake (at age six months) in recent years before the pandemic shifted to a decreasing trend after the pandemic started, which accelerated slightly post-pandemic. Similarly, the matched-sibling analysis found significantly lower odds of vaccination for children eligible during and even more for children after the pandemic compared to their pre-pandemic siblings in households with both at least one vaccinated and an unvaccinated child. Both analyses indicate a negative effect of the pandemic on first-dose DTaP-IPV uptake, with a sustained decline post-pandemic. This sustained decrease may reflect lingering pandemic effects or new post-pandemic factors, and is concerning for increased risk of vaccine-preventable disease outbreaks and weakened herd immunity.

The ITS analysis also showed level changes at both predefined interruptions. The abrupt decline in vaccination uptake at the start of the pandemic is most likely due to disruptions because of the COVID-19 pandemic and the public health measures in place. This was previously reported for MMR1 vaccination in the Netherlands [3]. The second interruption at the end of the pandemic coincided with the introduction of informed consent for vaccination registration. Although the estimated level change from our model (−1.00 percentage points in the unadjusted model, −0.66 percentage points in the adjusted model) is a bit smaller than the estimated proportion of vaccinated individuals for whom informed consent for registration in Praeventis is lacking (1.7%), the abrupt decline at this timepoint is most likely explained by this policy change. The matched-sibling analysis offers important additional insights beyond the ITS approach, as it inherently controls for family-level characteristics that remain constant between siblings. By comparing siblings within the same family, this method helps to isolate the independent effect of the pandemic from other potential confounding factors, including trends in the composition of the population.

Our findings are consistent with evidence from other European countries that have reported reductions in childhood vaccination uptake during the pandemic, with coverage for key vaccines such as MMR and DTaP-IPV in several regions falling below the WHO target of 90% [15-18]. In most cases, only partial recovery has been observed post-pandemic, and persistent declines have been accompanied by widening socioeconomic disparities [17, 18]. These similarities suggest that the negative impact of the pandemic on vaccination uptake is a broader phenomenon within Europe. However, it should be noted that most studies were conducted relatively soon after the initial pandemic period, which limits abilities to draw conclusions about longer-term impact on vaccination uptake. Outside of Europe, similar effects have been reported, although local factors such as healthcare system disruptions may influence the magnitude and duration of impact [19, 20]. These findings underline the importance of ongoing monitoring of vaccination uptake and initiating efforts to further study and address lower vaccination rates post-pandemic.

The sustained decrease in vaccination uptake observed post-pandemic suggests that lingering pandemic effects play a role or there are new post-pandemic reasons. A frequently mentioned result of the pandemic is a decrease in vaccine confidence [6]. In Flanders (Belgium), a cross-sectional survey found that, although most parents remained confident in childhood vaccines, overall confidence declined over time [21]. UK-based studies reported that, while overall confidence in routine childhood vaccination remains relatively high, it has declined in the post-pandemic era [22-24]. Particularly among ethnic minority groups, parents had more questions about childhood vaccinations compared to pre-pandemically [22]. A systematic review further supports that the pandemic affected vaccine confidence in various countries, though the direction and extent of change are context-dependent and may evolve over time [25]. In the Netherlands, parental vaccine confidence appears to have slightly decreased after the pandemic [7], but more specific reasons remain insufficiently understood and require further research.

Several factors may have contributed to this observed decrease in vaccine confidence following the COVID-19 pandemic. The widespread circulation of misinformation and disinformation during the pandemic, particularly through social media and other online platforms, likely played a significant role in shaping negative attitudes towards vaccination [5, 26-28]. Furthermore, public trust in governmental institutions was affected by the rapidly evolving situation and restrictive measures, while the fast development and rollout of COVID-19 vaccines raised concerns about safety and regulation [5]. Together, these factors may have undermined overall confidence in established vaccination programmes and contributed to the sustained decline in routine childhood vaccine uptake, although further research is needed to confirm this.

Several studies have previously examined the association between birth order and childhood vaccine uptake, with evidence suggesting that later-born children are at increased risk of incomplete or delayed immunization compared to first-borns, possibly due to factors such a parental time constraints or changing perceptions with subsequent children [11, 29]. This is in line with our finding that second-born children are less likely to be vaccinated and is concerning because, if this pattern of declining vaccine uptake over time persists, vaccination coverage could decline even further in the future if families have more children.

A key strength of this study is the use of the two complementary methodological approaches, enhancing validity and interpretability of our findings by providing both a broad epidemiological perspective and a more refined within-family comparison. Notably, both analyses consistently indicated a negative effect since the pandemic on DTaP-IPV vaccine uptake, lending greater confidence to the observed association. Furthermore, the use of nationwide, registry-based data enabled a comprehensive assessment of vaccination trends and adjustment for key sociodemographic factors. Lastly, by using recent data extending through 2025, we were able to evaluate both immediate and longer-term effects after the COVID-19 pandemic on childhood vaccination uptake, in contrast to earlier studies that have largely been limited to short-term outcomes.

Several limitations should be considered. First, post-pandemic vaccination coverage is likely underestimated due to incomplete registration following the introduction of informed consent; for primary DTaP-IPV, the average percentage of parents not providing consent is relatively low (1.7% compared to compared to over 3–10% for DTP booster and school-age vaccinations)[4]. Although sensitivity analyses continued to show a negative effect, variation in consent rates across subgroups may still have introduced bias, which we were unable to adjust for due to lack of data. Second, while the ITS controlled for pre-existing trends, unmeasured confounding remains possible given concurrent pandemic-related factors that may have influenced uptake. In addition, sociodemographic variables were measured at a single time point (2024) and may not reflect changes over the study period, though major shifts are unlikely. Third, children who died before 31 December 2024 were excluded, slightly underrepresenting the at-risk population, but the impact is minimal given low childhood mortality. Fourth, some children may have received their vaccination later than usual due to pandemic delays, but assessing vaccination status at 9 months of age showed only a minor increase in coverage (0.2 percentage points) and no differences between pandemic periods (data not shown). This confirms that although there was some delay as described earlier, this was not substantial [3]. Finally, the matched-sibling analysis estimates effects only within families where siblings differ in both vaccination status and pandemic period, rather than across the entire population, which may limit the generalizability of the findings [12]. Generalizability of covariates associated with lower vaccination coverage may not hold true for one-child families, and the results could be influenced by unmeasured confounding, such as changes in family circumstances (e.g., divorce or household composition). Nonetheless, only 5% of households in this analysis included siblings who did not share the same parents (0.4% different mother; 4.6% different father), and excluding these households yielded nearly identical results (data not shown). Similarly, restricting the analysis to families with exactly two children produced similar findings, further supporting the robustness of our results.

## Conclusion

This nationwide study demonstrates a sustained decline in routine childhood vaccination uptake in the Netherlands during the COVID-19 pandemic that continued after the pandemic. Both population-level and within-family analyses indicate that the pandemic has had a negative impact on parental vaccination decisions, which may reflect lingering pandemic effects or new post-pandemic factors. The observation that the downward trend in vaccination coverage continues, rather than stabilizes, is particularly concerning as it may increase the risk of outbreaks of vaccine-preventable diseases and undermine herd immunity. Therefore, continuous monitoring of vaccination uptake and further research is essential to understand changes in practical barriers to and attitudes towards vaccination, alongside continued efforts to manage mis- and disinformation and address vaccine hesitancy.

## Data Availability

All data is available within CBS Microdata and can be made available under strict conditions.

