## Supplementary material for "Assessing the impact of the COVID-19 pandemic on routine childhood vaccination uptake in the Netherlands"

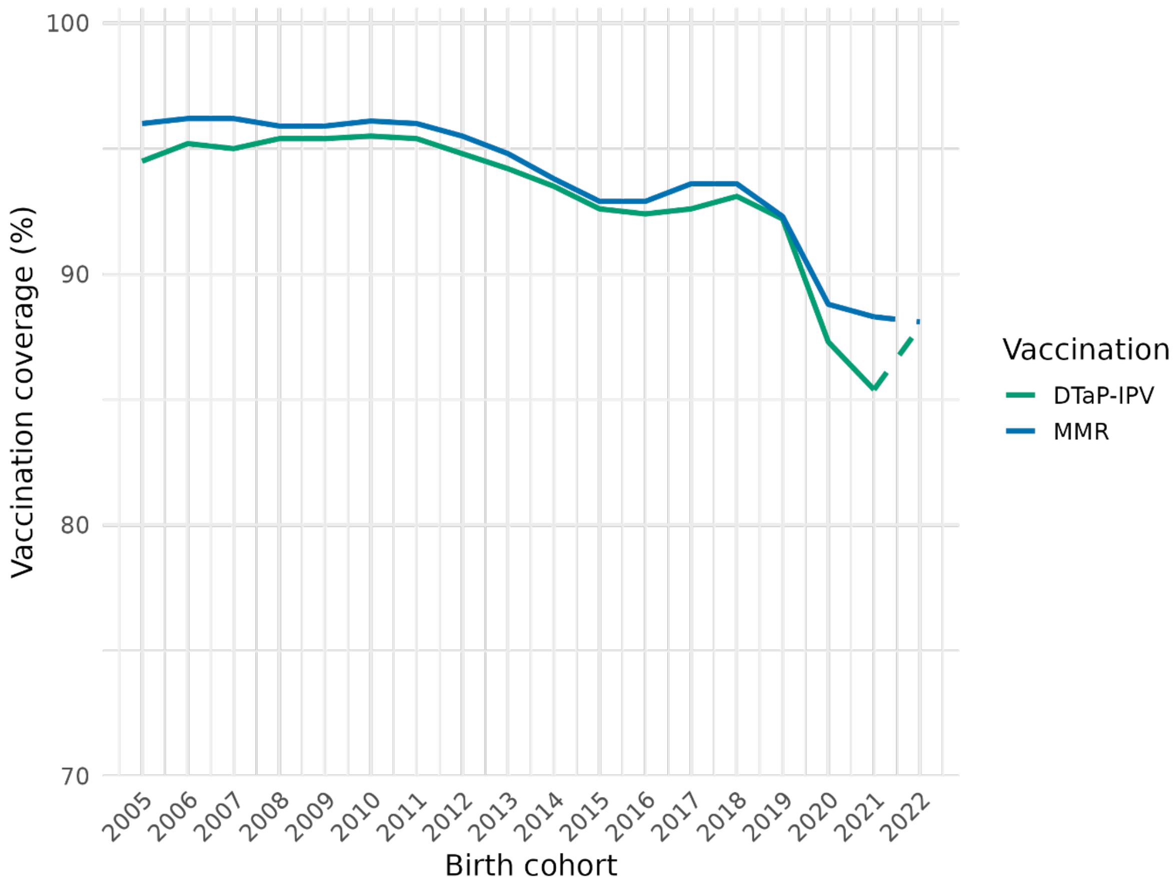


**Figure S1. First-dose MMR and ≥ 3 dose DTaP-IPV vaccination coverage at age 2 years, birth cohort 2005-2022, The Netherlands. Data retrieved from the annual vaccination coverage report by the Dutch National Public Health Institute [4].**

Abbreviations: *DTaP-IPV* diphtheria-tetanus-pertussis-polio, *MMR* measles-mumps-rubella

The dashed lines indicate the vaccination coverage is impacted by the informed consent. Note: changes in registered vaccination coverage in the most recent cohort do not necessarily reflect changes in actual coverage, as recent figures do not yet account for anonymous vaccinations or administrative corrections for missing JGZ indications. For DTaP-IPV, the apparent increase in coverage in cohort 2025 is due to such an administrative correction and does not indicate a true rise in uptake; actual coverage is likely slightly lower than the previous yea

**Table S2. Sociodemographic variables definitions**

| Variable | Definition | Levels | Reference year |
|---|---|---|---|
| Migration status | Migrations status based on whether:<br>1) children were born in the Netherlands,<br>2) parents were born in the Netherlands and<br>3) grandparents were born in the Netherlands | Dutch origin; first-generation migrant; child of first-generation migrant; child of second-generation migrants | 2024 |
| Country of origin | Country of origin was primarily defined by the country of birth (CoB) of the child itself. If the child was born in the Netherlands, country of origin was first determined by the parents' country of birth (CoB). If both parents were born in the Netherlands, the grandparents' CoB was then considered. Within this definition, the mother's CoB was prioritized over the father's, and the grandmother's CoB was prioritized over the grandfather's. Foreign-born origins were given precedence over Dutch origin if present in the lineage. | The Netherlands; Europe (excluding the Netherlands); Morocco; Turkey; Surinam; Dutch Caribbean; Indonesia; Other, America/Oceania; Other, Africa; Other, Asia). Indonesia; Other (Africa, Asia, America/Oceania) | 2024 |
| Maternal education level | Highest level of completed education.<br>High:<br>HBO higher professional education;<br>WO, University education<br>Not high:<br>primary education; VMBO, pre-vocational education;<br>MBO1, entry-level secondary vocational education; | High; not high | 2024 |

| | | | |
|---|---|---|---|
| | HAVO, senior general secondary education<br>WO, pre-university education | | |
| Household standardised disposable income | Disposable income of the household taking into account differences in household size and household composition, divided into quartiles based on the entire Dutch population. | First quartile; second quartile; third quartile; fourth quartile; unknown | 2024 |
| Level of Urbanisation | Urbanisation level was defined based on the average address density per square kilometre: extremely urbanised (≥2500 addresses/km2), strongly urbanised (1500-2499 addresses/km2), moderately urbanised (1000-1499 addresses/km2), hardly urbanised (500-999 addresses/km2) and not urbanised (≤500 addresses/km2). | Non-urbanised; hardly urbanised; moderately urbanised; strongly urbanised; extremely urbanised; unknown | 2024 |

**Table S3. Total effects of birth order and pandemic period on first-dose DTaP-IPV vaccination uptake among children in families with at least two children (reference group: first-born child, pre-pandemic period)**

| Group (ref: first child pre-pandemic) | OR | 95%CI |
|---|---|---|
| Second child pandemic | 0.15 | 0.11-0.21 |
| Second child post pandemic | 0.06 | 0.04-0.08 |
| Third child pandemic | 0.08 | 0.03-0.20 |
| Third child post pandemic | 0.02 | 0.01-0.06 |
| Fourth or later child pandemic | 0.02 | 0.00-5.94 |
| Fourth or later child post pandemic | 0.01 | 0.00-3.68 |

**Table S4. Matched-sibling conditional logistic regression model for first-dose DTaP-IPV vaccination uptake at age 6 months, children born 2016-2024 within families with two children (n = 554,496)**

| | **OR** | **95% CI** | **p value** |
|---|---|---|---|
| Pre-pandemic (ref) | ref | ref | ref |
| Pandemic | 0.50 | 0.43-0.58 | <0.001 |
| Post pandemic | 0.14 | 0.12-0.18 | <0.001 |
| First born (ref) | ref | ref | ref |
| Second born | 0.32 | 0.29-0.35 | <0.001 |

**Table S5. Sensitivity analysis with 16% of unvaccinated children in the post-pandemic/informed consent period reclassified as vaccinated. Matched-sibling conditional logistic regression model for first-dose DTaP-IPV vaccination uptake at age 6 months, children born 2016–2024 within families with at least two children (n = 708,059)**

| | **Model 1** | | | **Model 2** | | |
|---|---|---|---|---|---|---|
| | OR | 95% CI | p value | OR | 95% CI | p value |
| Pre-pandemic (ref) | ref | ref | ref | ref | ref | ref |
| Pandemic | 0.43 | 0.40-0.47 | <0.001 | 0.48 | 0.43-0.54 | <0.001 |
| Post pandemic | 0.35 | 0.32-0.40 | <0.001 | 0.36 | 0.32-0.41 | <0.001 |
| First born (ref) | ref | ref | ref | ref | ref | ref |
| Second born | 0.49 | 0.46-0.52 | <0.001 | 0.53 | 0.47-0.58 | <0.001 |
| Third born | 0.34 | 0.30-0.39 | <0.001 | 0.31 | 0.19-0.51 | <0.001 |
| Fourth or later born | 0.40 | 0.32-0.50 | <0.001 | 0.21 | 0.01-4.62 | 0.320 |
| pandemic* second born | | | | 0.82 | 0.69-0.98 | 0.026 |
| post pandemic * second born | | | | 0.96 | 0.84-1.11 | 0.584 |
| pandemic * third born | | | | 0.90 | 0.53-1.51 | 0.683 |
| post pandemic * third born | | | | 1.14 | 0.69-1.87 | 0.616 |
| pandemic * fourth or later born | | | | 0.51 | 0.02-13.03 | 0.687 |
| post pandemic * fourth or later born | | | | 2.01 | 0.09-45.14 | 0.661 |

**Table S6. Total effects of birth order and pandemic period on first-dose DTaP-IPV vaccination uptake among children in families with at least two children (sensitivity analysis reclassifying 16% of unvaccinated children in the post-pandemic/informed consent period as vaccinated; reference group: first-born child, pre-pandemic period)**

| Group (ref: first child pre-pandemic) | OR | 95%CI |
|---|---|---|
| Second child pandemic | 0.21 | 0.11-0.21 |
| Second child post pandemic | 0.18 | 0.04-0.08 |
| Third child pandemic | 0.13 | 0.03-0.20 |
| Third child post pandemic | 0.13 | 0.01-0.06 |
| Fourth or later child pandemic | 0.05 | 0.00-5.94 |
| Fourth or later child post pandemic | 0.15 | 0.00-3.68 |
